# Quantifying spatial heterogeneity of malaria in the endemic Papua region of Indonesia: analysis of epidemiological surveillance data

**DOI:** 10.1101/2022.04.18.22273950

**Authors:** Ihsan Fadilah, Bimandra A. Djaafara, Karina D. Lestari, Sri Budi Fajariyani, Edi Sunandar, Billy Graham Makamur, Berry Wopari, Silas Mabui, Lenny L. Ekawati, Rahmat Sagara, Rosa N. Lina, Guntur Argana, Desriana E. Ginting, Maria Endang Sumiwi, Ferdinand J. Laihad, Ivo Mueller, Jodie McVernon, J. Kevin Baird, Henry Surendra, Iqbal R. F. Elyazar

## Abstract

**Background:** As control efforts progress towards elimination, malaria is likely to become more spatially concentrated in few local areas. The purpose of this study was to quantify and characterise spatial heterogeneity in malaria transmission-intensity across highly endemic Indonesian Papua.

**Methods:** We analysed individual-level malaria surveillance data for nearly half a million cases (2019–2020) reported in the Papua and West Papua provinces and adapted the Gini index approach to quantify spatial heterogeneity at the district and health-unit levels. We showed malaria incidence trends and the spatial and temporal distribution of sociodemographic characteristics and aetiological parasites among cases.

**Findings:** While Papua province accounted for the majority of malaria cases reported in the region and had seen a rise in transmission since 2015, West Papua province had maintained a comparatively low incidence. We observed that Gini index estimates were high, particularly when the lower spatial scale of health units was evaluated. The Gini index appears to be inversely associated to annual parasite-incidence, as well as the proportions of vivax malaria, male sex, and adults.

**Interpretation:** This study suggests that areas with varying levels of transmission-intensities exhibited distinct characteristics. Malaria was distributed in a markedly disproportionate manner throughout the region, emphasising the need for spatially targeted interventions. Periodic quantification and characterisation of risk heterogeneity at various spatial levels using routine malaria surveillance data may aid in tracking progress towards elimination and guiding evidence-informed prioritisation of resource allocation.

**Funding:** Strengthening Preparedness in the Asia-Pacific Region through Knowledge (SPARK) project.

**Research in context:** *Evidence before this study:* We searched PubMed up to and including November 19, 2021, for relevant articles on the spatial distribution of malaria in the Papua region of Indonesia, using the terms (“malaria”) AND (“distribution” OR “variation” OR “heterogeneity” OR “cluster” OR “aggregation”) AND (“Papua”) AND (“Indonesia”). Despite the region’s mostly stable transmission areas, there has been considerable variation in transmission intensity across the region. According to community surveys conducted up to 2010, estimates of parasite prevalence of *Plasmodium falciparum* and *Plasmodium vivax* were highly variable, ranging from 0% to at least 40% and from 0% to at least 7%, respectively, across the region. Similarly, when the Papuan subset of the 2007 National Basic Health Research data was used, the degree of spatial heterogeneity in malaria risk among Papuan districts remained apparent even after sociodemographic were adjusted. Current evidence that is more representative of the current situation, including an easily interpretable and comparable measure of spatial heterogeneity across space and time, is limited.

*Added value of this study:* Our analysis of large-scale and routinely collected malaria surveillance data from January 2019 to December 2020 revealed significant spatial heterogeneity across the Papua region, as measured by the Gini index. Complementing conventional approaches using geospatial maps and risk tables, the Gini index can be used to provide a single, and sensitive numerical indicator summarising the degree of transmission heterogeneity at a specified spatial level of interest. Along with the previously recognised high spatial heterogeneity among districts, this study revealed a greater degree of intra-district heterogeneity at the health-unit level. That is, within the districts, there were also few health centres and hospitals with a disproportionately higher malaria burden. We observed distinct characteristics of individuals who contracted malaria in districts with varying levels of incidence. The higher transmission magnitude was associated with a lower Gini index, as well as with lower proportions of vivax malaria, male sex, and adults among the cases.

*Implications of all the available evidence:* This study provides contemporary empirical evidence for the spatial heterogeneity of malaria distribution across the Papua region of Indonesia, particularly at the lower spatial resolution of health units. Evaluating spatial heterogeneity at a lower spatial scale is likely essential to refine and update local malaria control strategic planning. The combination of comprehensive, routine malaria surveillance data and the Gini index may enable policymakers to assess the magnitude and characteristics of spatial heterogeneity with increased frequency, interpretability, and comparability, allowing for the rapid identification of transmission foci and the deployment of public health measures. Effective control of parasite reservoirs associated with intense transmission may further shrink the risk of infection in adjacent areas with a lower degree of malaria exposure.

## Background

Despite its declining trend, malaria continues to be a significant global health burden, with an estimated 229 million cases and 409,000 deaths in 2019.^1^ As transmission declines, malaria risk becomes more heterogeneous and is often concentrated in specific locations or populations. Heterogeneity of infection refers to the clustering of infection and clinical disease such that a small fraction of individuals bears most of the population’s burden.^2^ Transmission heterogeneity can be defined in terms of spatial or temporal boundaries, as well as individual- and environment-level characteristics purported to be disease determinants.

To better target interventions in areas moving towards elimination, it is critical to detect and quantify transmission heterogeneity and understand factors that may contribute to disease persistence in these locations.^3^ The detection of local level clusters of infection can have an important role in understanding the micro-epidemiological patterns of disease transmission, and ensuring that control strategies are tailored to the specific epidemiological characteristics in an area as much as is feasible.^4^ Mathematical modelling indicates that well-targeted interventions could have a four-fold impact on the population to untargeted interventions.^5^

Indonesia has been one of the major malaria hotspots in the WHO South-East Asia region, with an estimated 658,380 cases and 1170 deaths in 2019.^1^ As in many Southeast Asian settings, malaria elimination in Indonesia becomes more challenging with the presence of multi-species infections combined with the difficulty of identifying in which locations persistent transmission might be occurring.^6^ Although more than half of the districts in Indonesia were declared malaria-free by 2017,^7^ two provinces in the Papua region (West Papua and Papua) still combat high transmission of the infection. In 2020, both provinces contributed to nearly 90% of all nationally reported cases, which poses a challenge for achieving the nationwide elimination goal targeted by 2030.^8^ However, robust assessment of transmission heterogeneity in Indonesia has been focused on national and provincial level estimates, mainly due to the availability of data.^9,10^

Synthesising estimates of parasite prevalence from independently conducted community surveys across Indonesia (up to 2010), Elyazar and colleagues observed highly heterogeneous transmission in the Papua region despite its status as containing mostly stable transmission areas (annual parasite incidence [API] of 0·1 or more cases per 1000 population under surveillance).^9,10^ The parasite prevalence of *P. falciparum* and *P. vivax* varied across the region from 0% to at least 40% and from 0% and at least 7%, respectively.^9,10^ Based on data from the Papuan subset of the 2007 National Basic Health Research (n = 21,772), risk heterogeneity of clinical malaria was detectable across districts, even after controlling for sociodemographic characteristics.^11^ However, empirical evidence on transmission heterogeneity in the context of current endemicity is scarce and no single summary estimate for this heterogeneity has been provided in Indonesian Papua. This study quantified and characterised spatial heterogeneity in the Papua region using contemporary and large-scale malaria surveillance data.

## Methods

### Study design and data sources

#### Malaria surveillance

We obtained anonymised, individual-level data of 446,786 notified malaria cases in the Papua region from the electronic malaria surveillance information system (Sismal) database between January 2019 and December 2020. While the surveillance system was established just over a decade ago as part of Indonesia’s National Malaria Control Programme, it has steadily improved over time. Recently, individual case data have become available electronically for more detailed analysis.

The database contains detailed information about the demographic, geographical, clinical, and epidemiological characteristics of malaria cases. Passive case detection, defined as the detection of cases by patients presenting with their illness to healthcare services, comprised 90% or more of the methods for identifying cases. Microscopy, rapid diagnostic testing, or polymerase chain reaction were used to confirm each reported case. Informed consent from patients included in these anonymised data was waived due to the secondary nature of these routine surveillance data.

Missing diagnostic days were uniformly imputed by the first day of the month, in which case was notified. If the diagnosis month was missing, the case was not considered. Additionally, we excluded a small number of observations due to missing data in a few other variables. For instance, to describe the proportion of infecting species in 2020, 0·04% (89 out of 220,435) of the data in species variable missing. We assumed that such a degree of missingness was unlikely to result in significant bias.

#### Population-at-risk

We estimated the population at risk of contracting malaria under surveillance using annual projected population sizes. Statistics Indonesia (*Badan Pusat Statistik*) provides these projections at the administrative level specified, e.g., provinces, districts (*kota/kabupaten*), either overall or by 5-year age group and sex.^12^ At the health unit level, catchment populations were generally undocumented. We divided the population-at-risk in a district by the number of within-district health units as recorded in the database or as officially published by the Ministry of Health for 2020,^13^ whichever was greater. Discrepancies in the numbers of health units between 2019 and 2020 were determined to be negligible, if they existed at all. For weekly or monthly calculations involving populations-at-risk, we assumed that the population-at-risk estimates were identical to the corresponding annual estimates, implying that the figures remained relatively stable over a given year.

### Data analysis

#### Transmission intensity and epidemiological characteristics

To estimate transmission intensity at the provincial, district, or health-unit level, we divided the number of laboratory-confirmed malaria cases (numerator) over a specified period (e.g., a week, month, or year) by the population-at-risk (denominator), expressed as cases per 1,000 population under surveillance. When the numerator is the total number of cases reported during a given year, the estimated incidence risk is commonly referred to as the annual parasite incidence per 1,000 (API per 1,000, or for simplicity, API).

We calculated malaria incidence risk by location, time period, age group, sex, and/or parasite aetiology. The proportions of cases are presented for various types of health units and occupation. Additionally, we used locally estimated scatterplot smoothing (loess) to visualise the relationship between the natural logarithm of API and district-level proportions of *P. vivax*, adults, and males, by year.

#### Quantifying spatial heterogeneity

We adapted the Gini index as a proxy for the spatially asymmetric distribution of transmission intensity across the Papua region. More than a century ago, Corrado Gini introduced this statistical measure to quantify the degree of wealth or income inequality within a specified population.^14^

Although it is most commonly used in economics, the Gini index has been widely applied in a diverse range of research, including infectious-disease epidemiology.^15^

As Lana and colleagues recently demonstrated in measuring the unequal distribution of malaria in Brazil,^16^ the Gini index *G* is defined by the formula

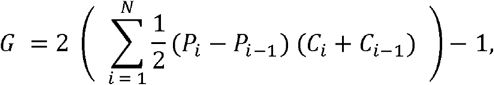

where *i* denotes the index for each spatial unit of interest (e.g., district, health unit), *N* is the number of spatial units at a given level (e.g., number of districts in a given province), *P*_*i*_ is the cumulative proportion of the population in unit *i, C*_*i*_ is the cumulative proportion of cases in unit *i*, and *P*_0_, *C*_0_ = 0. The formula is evaluated by first ranking the units in decreasing order of incidence risk (i.e., API). The Gini index is a numeric value between 0 and 1 (equivalently, 0% and 100%), where *G* = 0 indicates a perfectly proportional distribution of cases to unit population sizes and *G* = 1 indicates a perfectly disproportionate distribution of cases in which only one unit is responsible for all the cases occurring in the region. The index was calculated at two levels, namely, districts and health units, for each province and district. The 95% confidence interval (95% CI) around the Gini index estimate was constructed using a data-driven, nonparametric percentile bootstrap with 10,000 replicates.^17,18^ To approximate the sampling distribution, we used an identical estimator for *G* in each bootstrap replicate. We employed linear regression to visualise the association between the within-district Gini index and the natural logarithm of API.

To aid interpretation, we visualised spatial heterogeneity using the Lorenz curve,^19^ where the vertical and horizontal axes correspond to the cumulative proportions of malaria cases and population (ranked from the lowest to highest API), respectively. The 45° diagonal line relates to a perfectly proportional distribution of cases in the region (*G* = 0). Deviations from the line imply some degree of spatial heterogeneity, where a larger area under the Lorenz curve corresponds to a Gini index closer to unity. We conducted all statistical analyses and visualised data using R version 4.1.0.

## Results

### Transmission intensity and epidemiological characteristics

Over the past decade (2011–2020), there were 2,046,833 reported malaria cases in Indonesian Papua. Of which, 1,820,566 (88·9%) and 226,267 (11·1%) cases were detected in Papua and West Papua, respectively. *P. falciparum* and *P. vivax*, either mono- or mixed-infections, caused 90–95% of these cases. The remaining small proportions were due to *P. malariae, P. ovale*, and aetiologically unspecified cases. Over the first half of the decade, the cumulative incidence of malaria decreased significantly (figure 1A). While incidence in West Papua continued to decline until the end of 2016, Papua experienced a rebound in 2015. For the remainder of the decade, Papua’s cumulative incidence increased steadily. In contrast, West Papua has maintained a low notified cases of less than two per 1000 population in recent years. In the last two years (2019–2020), we observed around 40% of cases in Papua were consistently caused by *P. vivax* (figure 1B). In West Papua, the share was initially higher of approximately 50% in 2019 but appears to have declined to around 40% in the following year.

**Figure 1.**
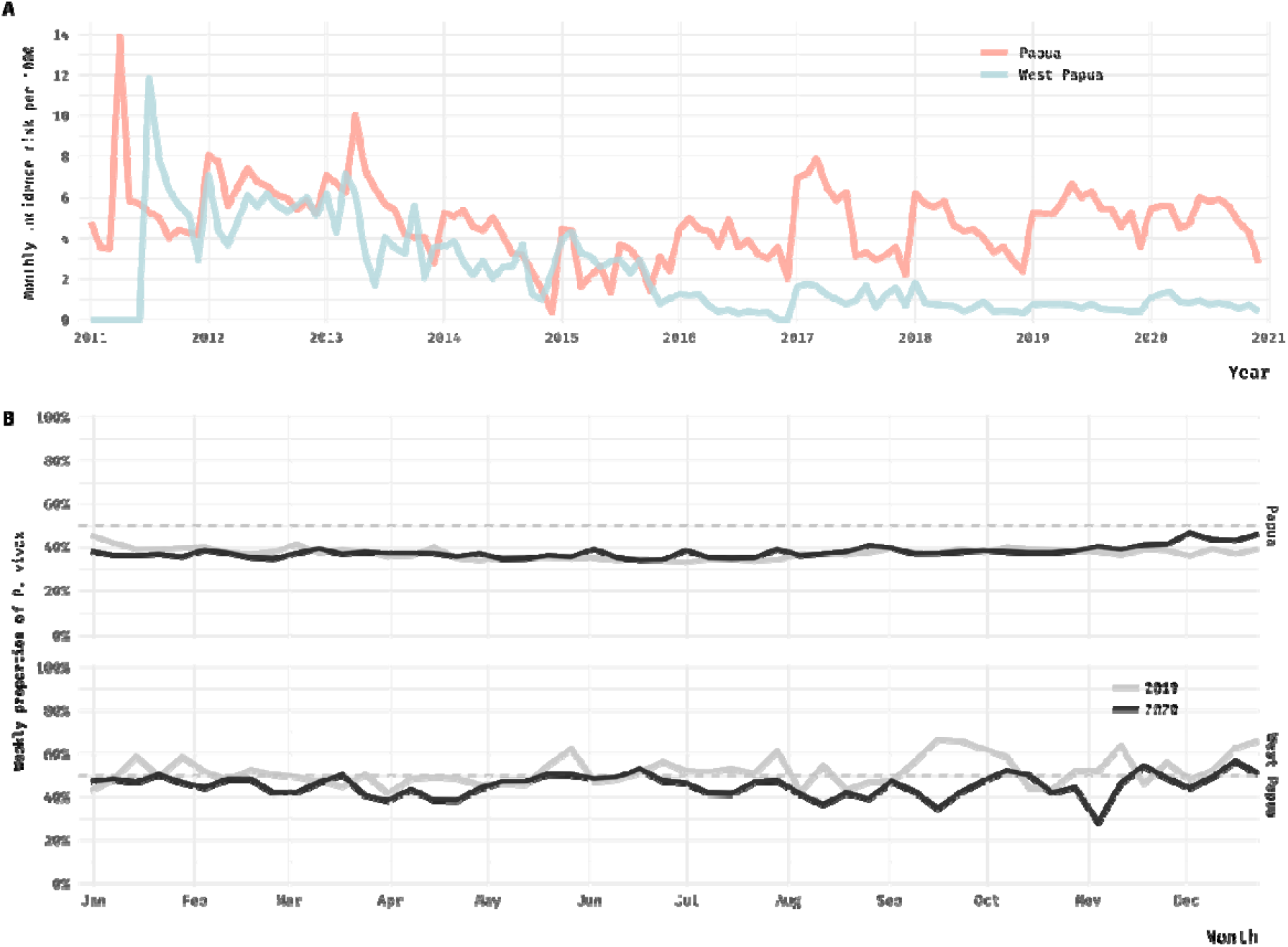
Malaria epidemiological trends in the Papua region, Indonesia. (A) Monthly incidence risk of malaria per 1,000 population in Papua and West Papua over the last decade (2011–2020). (B) Weekly proportions of *P. vivax* malaria infections in 2019 and 2020. The dashed horizontal line indicates when *P. vivax* and *P. falciparum* had been present in equal proportions in all cases.

The annual parasite incidence varied widely across the region (figures 2A, 2B, and S1). In a few districts (e.g., Keerom, Mimika, Sarmi), API reached nearly 400 cases per 1000 population. In other districts, such as Lanny Jaya, Sorong Selatan, or Maybrat, it was less than one case per 1000 population. Of the 42 districts across the region during 2020, seven (16·7%), ten (23·8%), 14 (33·3%), and 11 (26·2%) had an API of [100, 1000), [10, 100), [1, 10), and [0, 1) cases per 1000 population, respectively. More districts with a lower API (e.g., API of [0, 10)) experienced a relative reduction in API than districts with a higher API (e.g., API of [10, 1000)) (figure S1).

**Figure 2.**
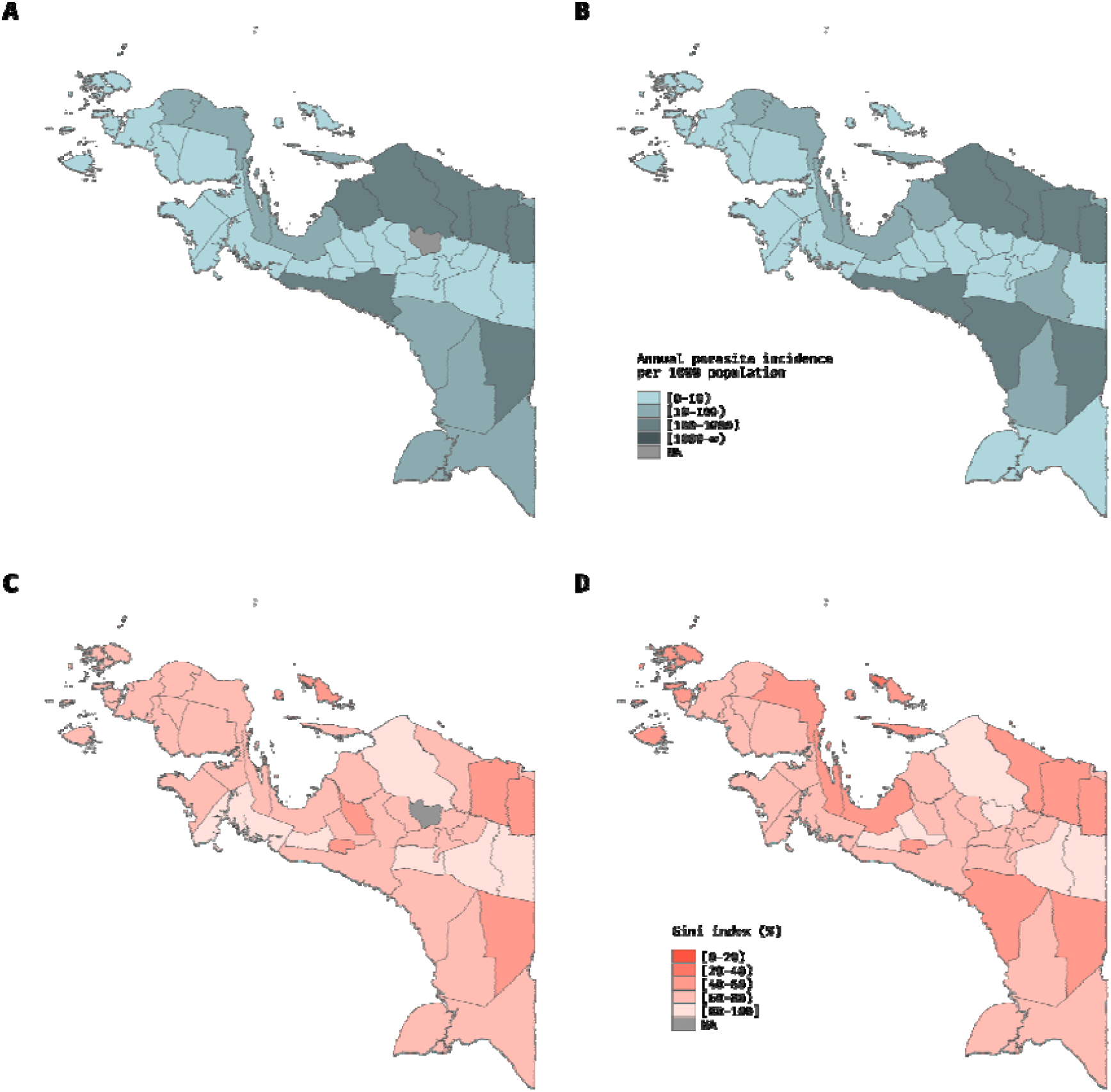
Spatially heterogeneous transmission intensity of malaria across districts in the Papua region, Indonesia. The polygon represents a district that has been coloured proportionally to the annual parasite incidence in 2019 (A) or 2020 (B), as well as the Gini index at health unit level in 2019 (C) or 2020 (D). Interval notation is used to represent categories of API and the Gini index.

Throughout 2019–2020, more males contracted malaria both in Papua (56–57%) and West Papua (59–63%) (figure 3A). API was highest in under-5 and young children, then gradually decreased in productive age and older groups (2020, figure 3B). This age-related pattern was not dissimilar in the preceding year of 2019. In terms of aetiology, *P. falciparum* and *P. vivax* were equally likely to cause malaria in West Papua in 2019, while *P. falciparum* caused a greater proportion of malaria in Papua. *P. falciparum* became more predominant in West Papua in 2020.

**Figure 3.**
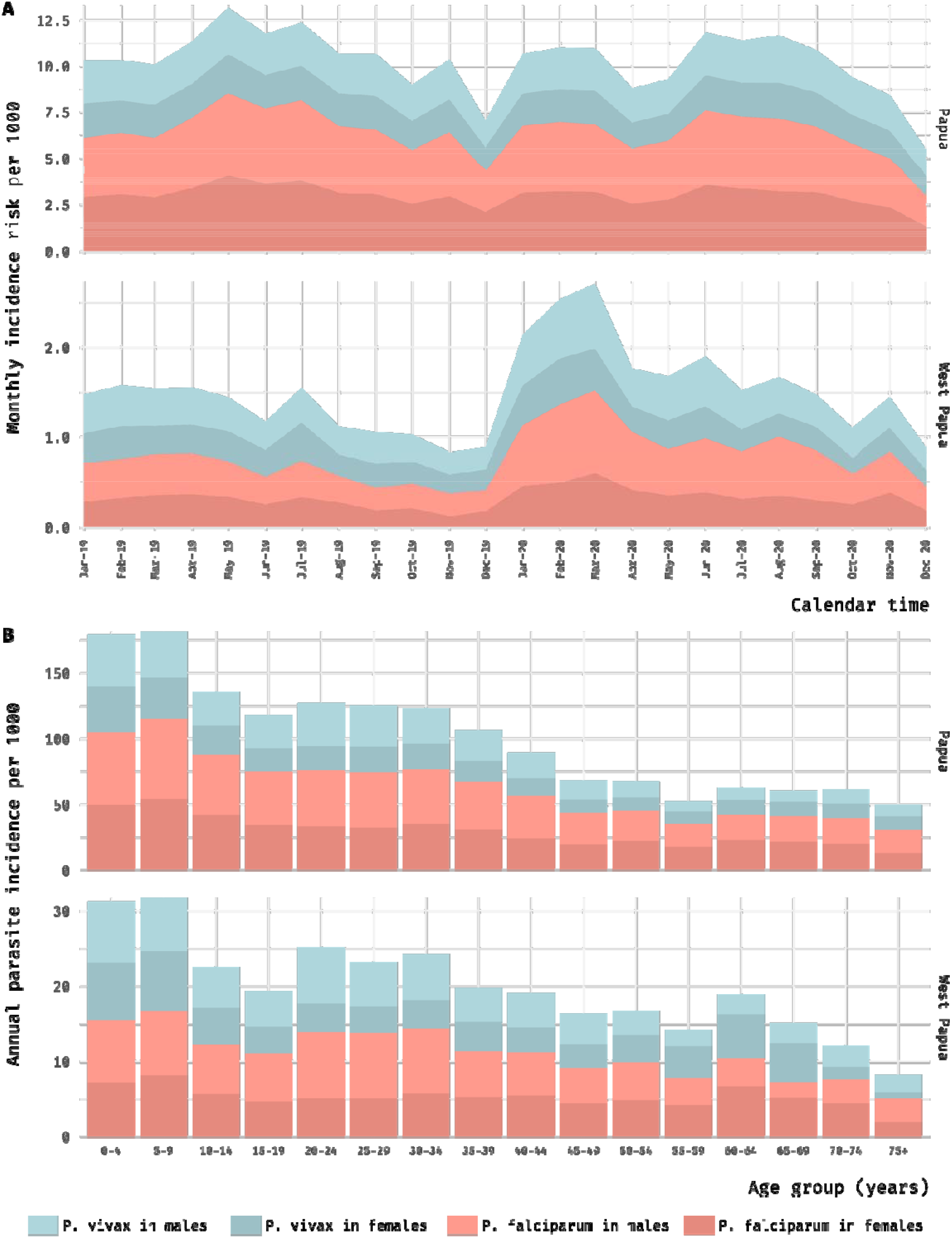
Distributions of malaria cases by age, sex, and parasite species in Papua and West Papua, Indonesia. (A) Monthly incidence risk of malaria per 1,000 population (2019–2020), by sex and species. (B) Annual parasite incidence by age group in 2020.

At the district level, for instance in three selected districts representative of a comparatively high, moderate, and low level of transmission intensity (Keerom, Nabire, Jayawijaya, respectively), the age patterns of cases in different settings appear distinctive (figure 4). We observed that API was highest in children in the high transmission setting (Keerom) but was particularly highest in young adults and productive age groups in the low transmission setting (Jayawijaya). Malaria events in Jayawijaya were predominated by *P. vivax*, in contrast to those in the moderate-to-high risk areas of Keerom and Nabire where *P. falciparum* remained the primary causative agent (figures 4 and S2). In the more moderate setting (Nabire), the age patterns were between the two extremes. Here, children under-5 years up to early-40 years adults appear to have approximately comparable API.

**Figure 4.**
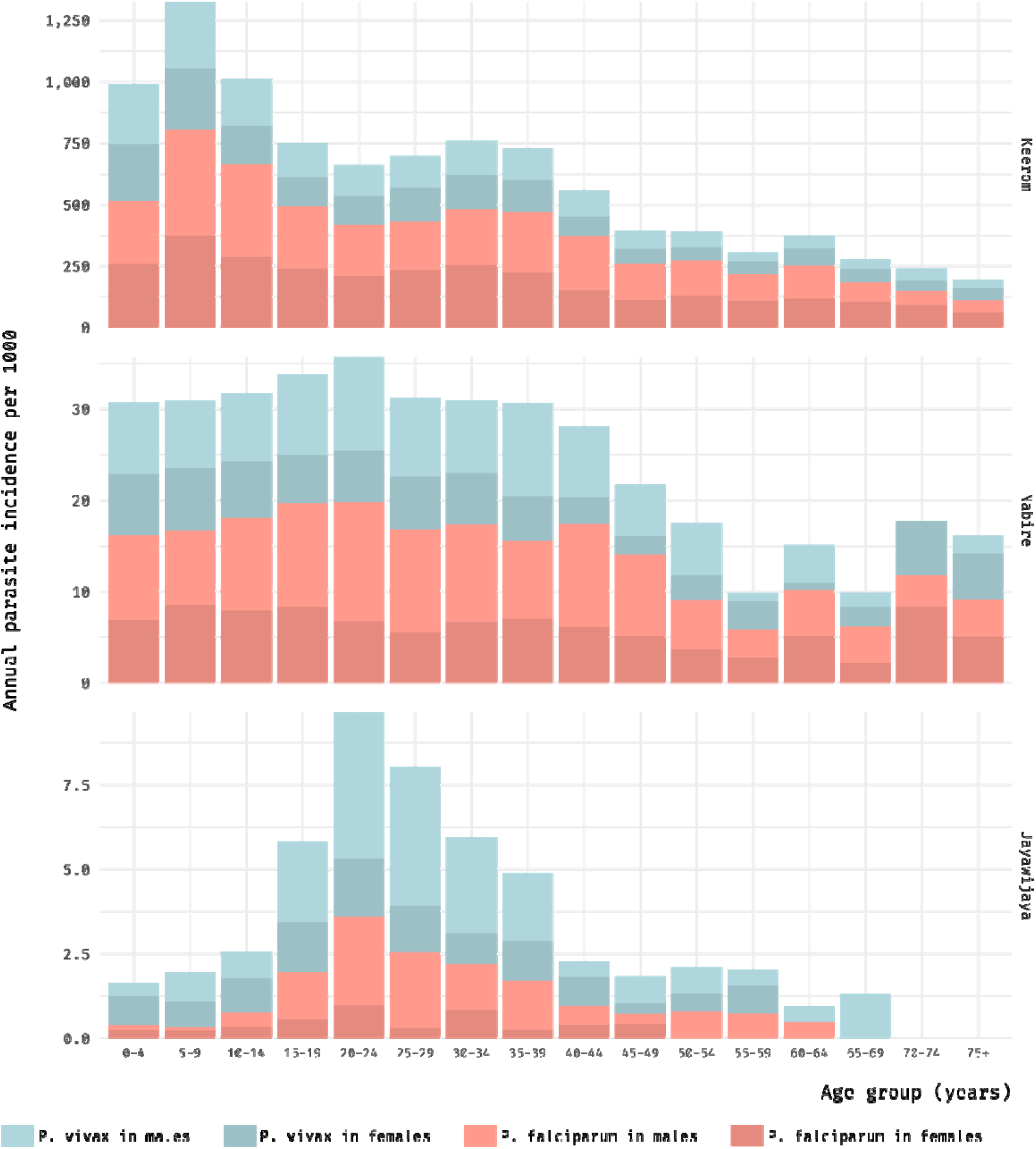
District-specific annual parasite incidence by age group in Keerom, Nabire, and Jayawijaya (2020). The plot illustrates a district that is representative of an area with a high level of transmission intensity (Keerom), a moderate level of transmission intensity (Nabire), and a low level of transmission intensity (Jayawijaya).

We found that parasite species, age, and sex of the notified cases were all associated with the magnitude of API in 2019 or 2020. Districts with a lower proportion of *P. vivax* tended to be in a higher transmission setting (figure 5A), implying the predominance of *P. falciparum*. Districts with a higher API were more likely to detect cases among younger demographics (figure 5B). For sex, we observed a similar pattern. As API increased, cases were more equally likely to occur between males and females (figure 5C). Males were most likely to develop cases in districts with a low API. Additionally, hospitals, which reported relatively more severe cases (figure S3), accounted for a sizeable proportion of malaria cases in some districts, including Mamberamo Raya and Merauke (figure S4). Among individuals aged 18 years and older, farmers and housewives formed a large majority of patients in most districts (figures 6A, 6B).

**Figure 5.**
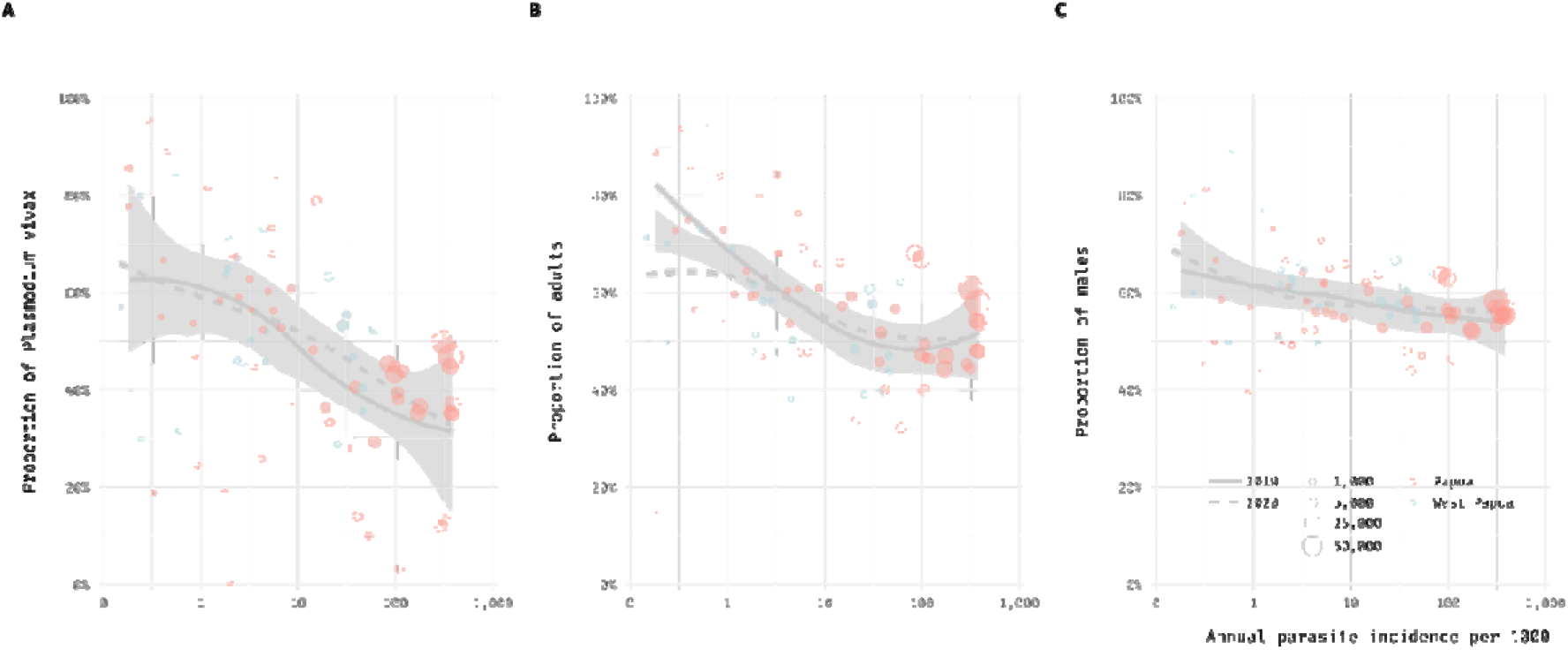
District-specific transmission intensity and its relation to the proportions of *P. vivax*, adults, and male sex among the notified cases. Proportions of (A) *P. vivax*, (B) adults (18 years and older), and (C) male sex against the natural logarithm of API. Smoothing of scatterplots was used to model the curves (loess). Shaded grey areas around the curves represent the 95% confidence intervals. Each dot is proportional in size to the total number of cases. Horizontal axes are represented by a logarithmic scale.

**Figure 6A.**
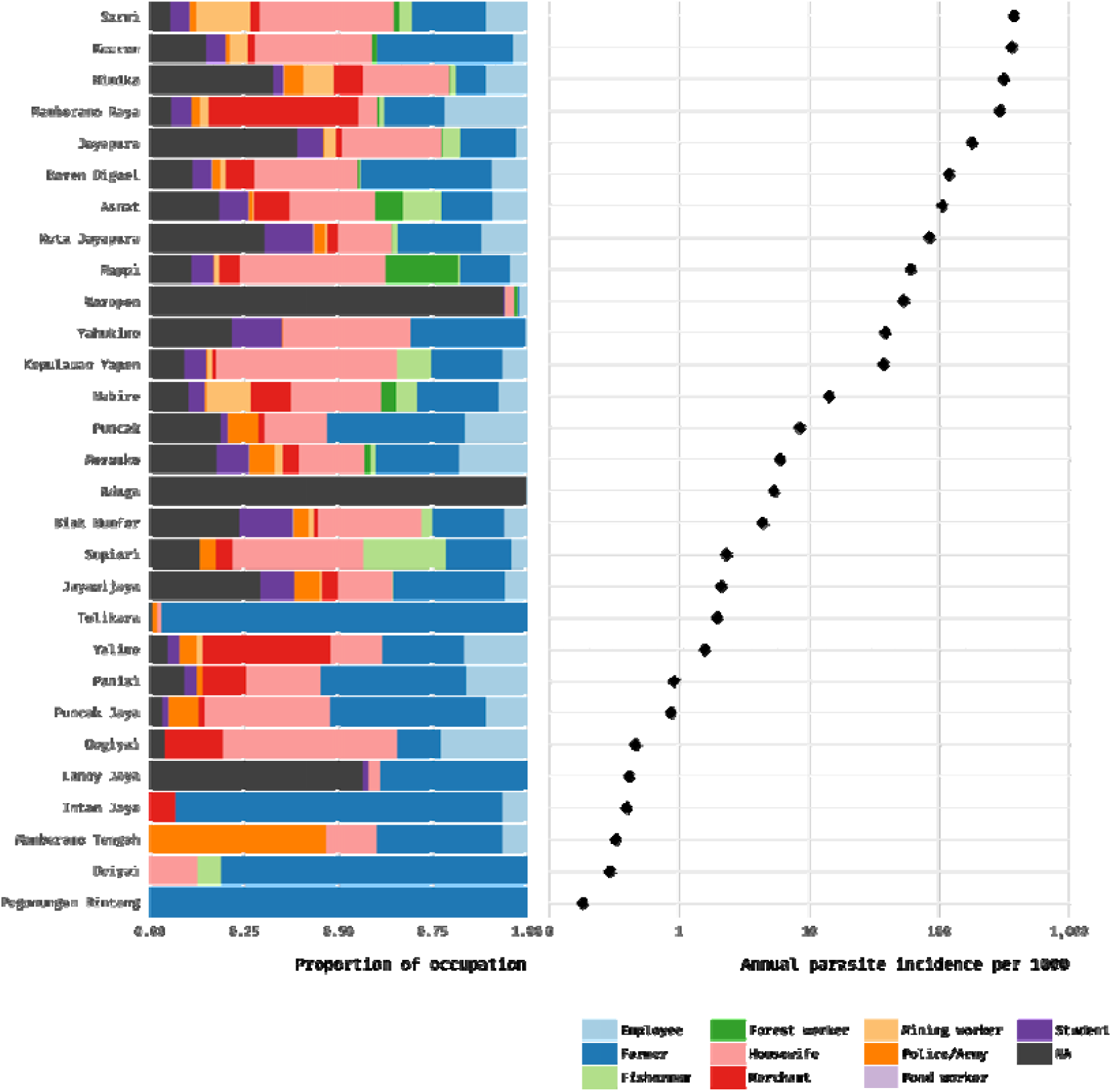
District-specific relative proportions of different occupation types in Papua (2020). Districts are arranged in decreasing order of API. Horizontal axis of the right-hand plot is shown on the logarithmic scale. NA not available.

**Figure 6B.**
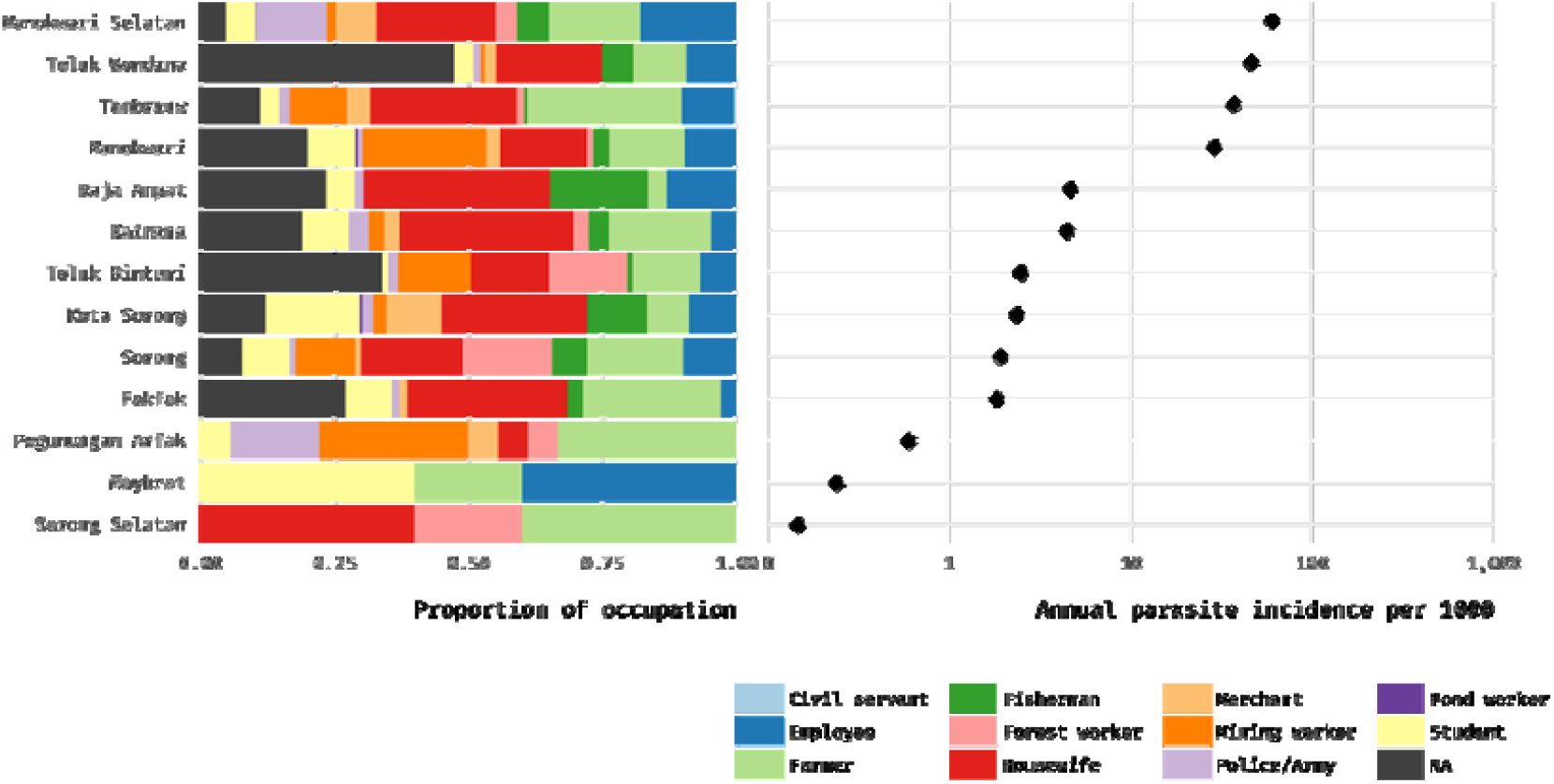
District-specific relative proportions of different occupation types in West Papua (2020). Districts are arranged in decreasing order of API. Horizontal axis of the right-hand plot is shown on the logarithmic scale. NA not available.

### Spatial heterogeneity of transmission intensity

The Lorenz curves represent the degree of spatial heterogeneity at two administrative levels (districts and health units) (figure 7). We found that all the curves deviated significantly from the 45° diagonal lines. At the district level, Papua Province consistently had a larger area under the curve than West Papua in both 2019 and 2020 (figure 7A). A larger area under the curve indicates a Gini index closer to one, or equivalently, 100% (table). Additionally, the curve provides an alternate perspective on heterogeneity. Approximately 80% of *P. falciparum* cases in 2020 occurred in districts that collectively accounted for about 20% of the population of Papua province.

**Figure 7.**
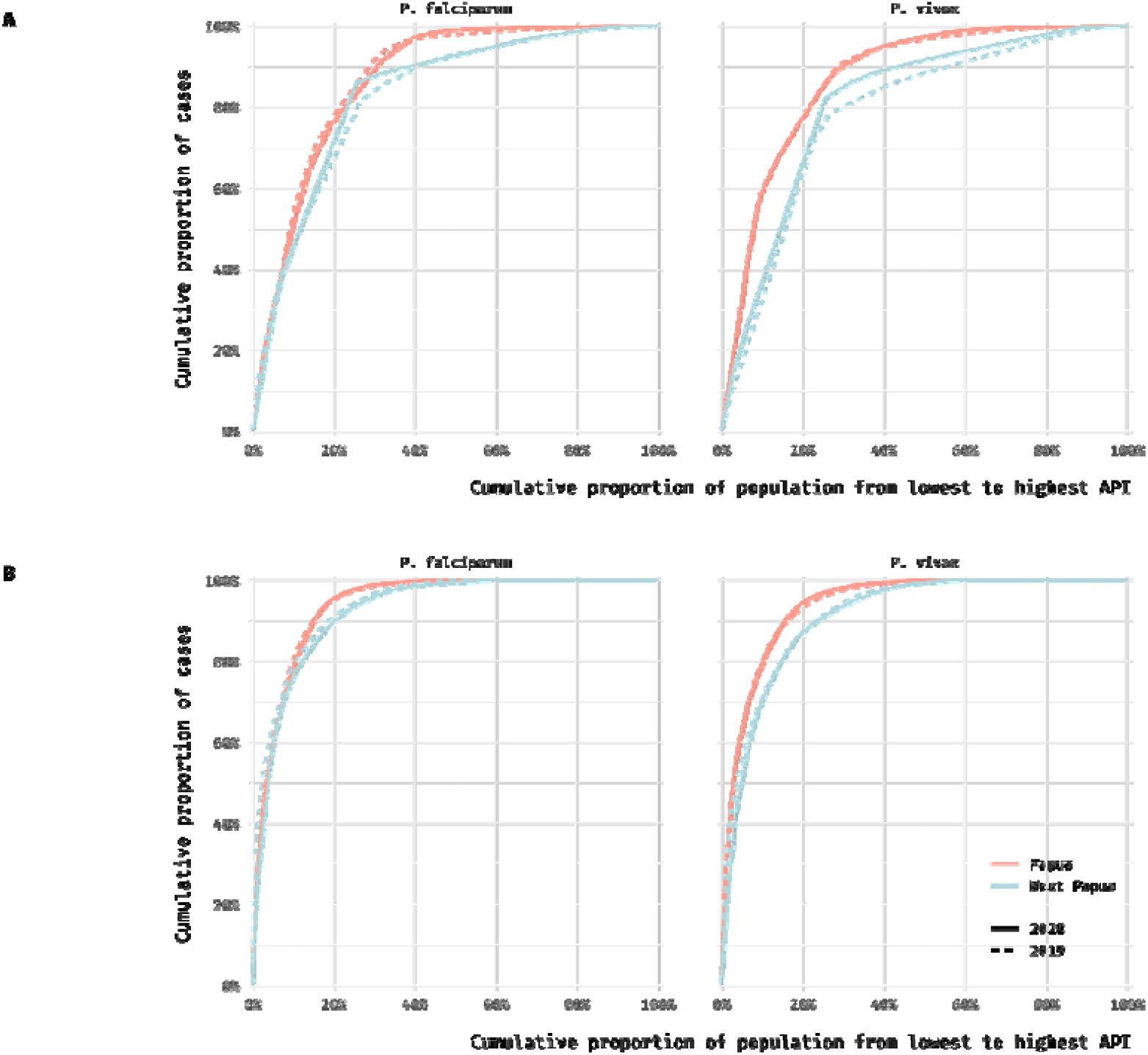
Lorenz curves for the spatial heterogeneity of transmission intensity, by species and administrative level. (A) District level. (B) Health unit level. Degree of heterogeneity is proportional to the area under the curve.

**Table.**
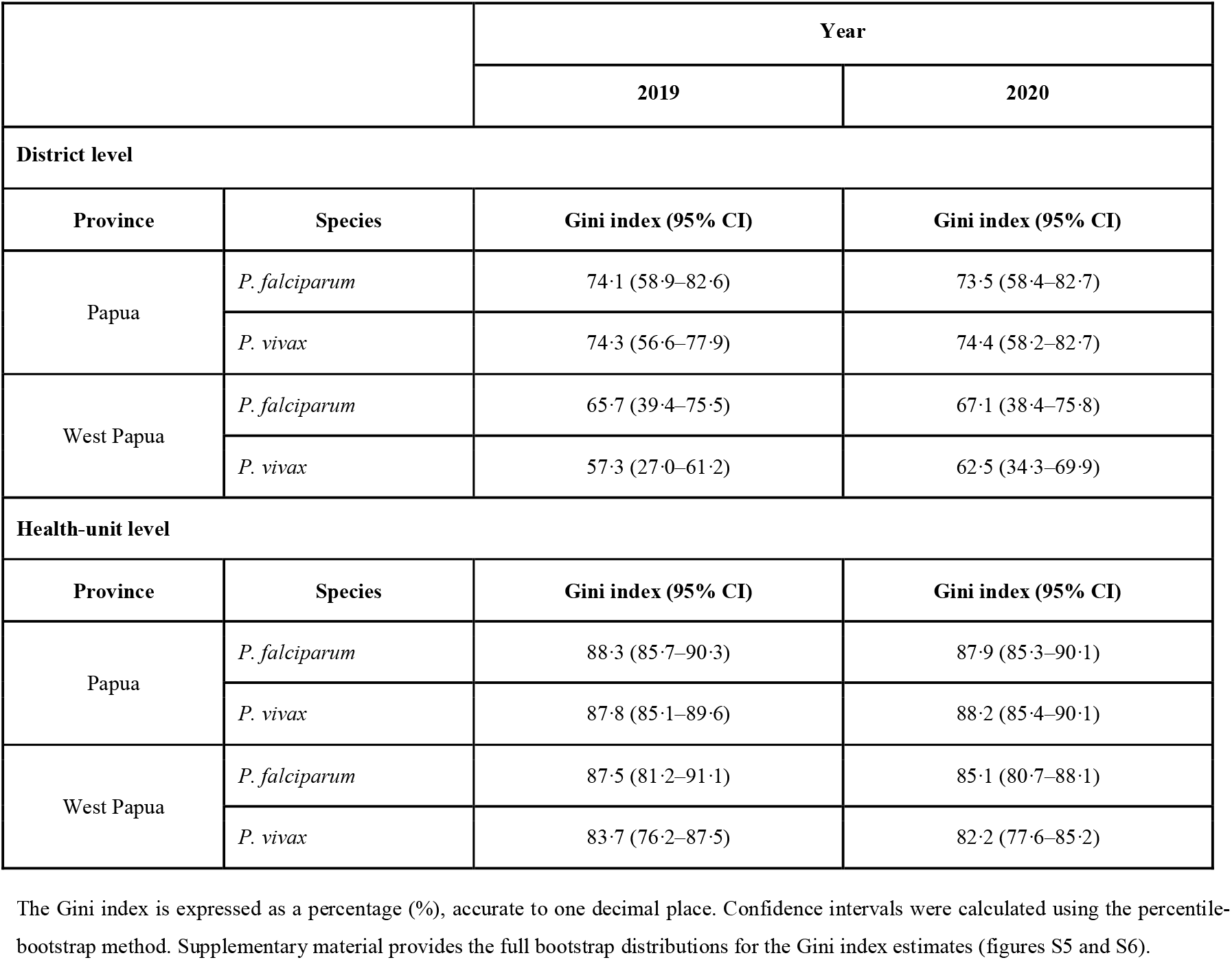
Gini index estimates for district and health unit levels, by year, province, and parasite species.

Assessing at the health unit level, we observed larger areas under the curves than at the district level (figure 7B), corresponding to a Gini index of more than 80% regardless of province, species, or year (table). At this level, for instance, approximately 80% of *P. falciparum* cases in 2020 occurred in districts that combined contained only 10% of the population size of Papua province.

We observed that even within-district Gini index estimates varied markedly across the Papua region (figures 2C and 2D). These estimates were evaluated at health unit level. Districts with a higher Gini index were more likely to be in a lower transmission setting and *vice versa* (figure 8).

**Figure 8.**
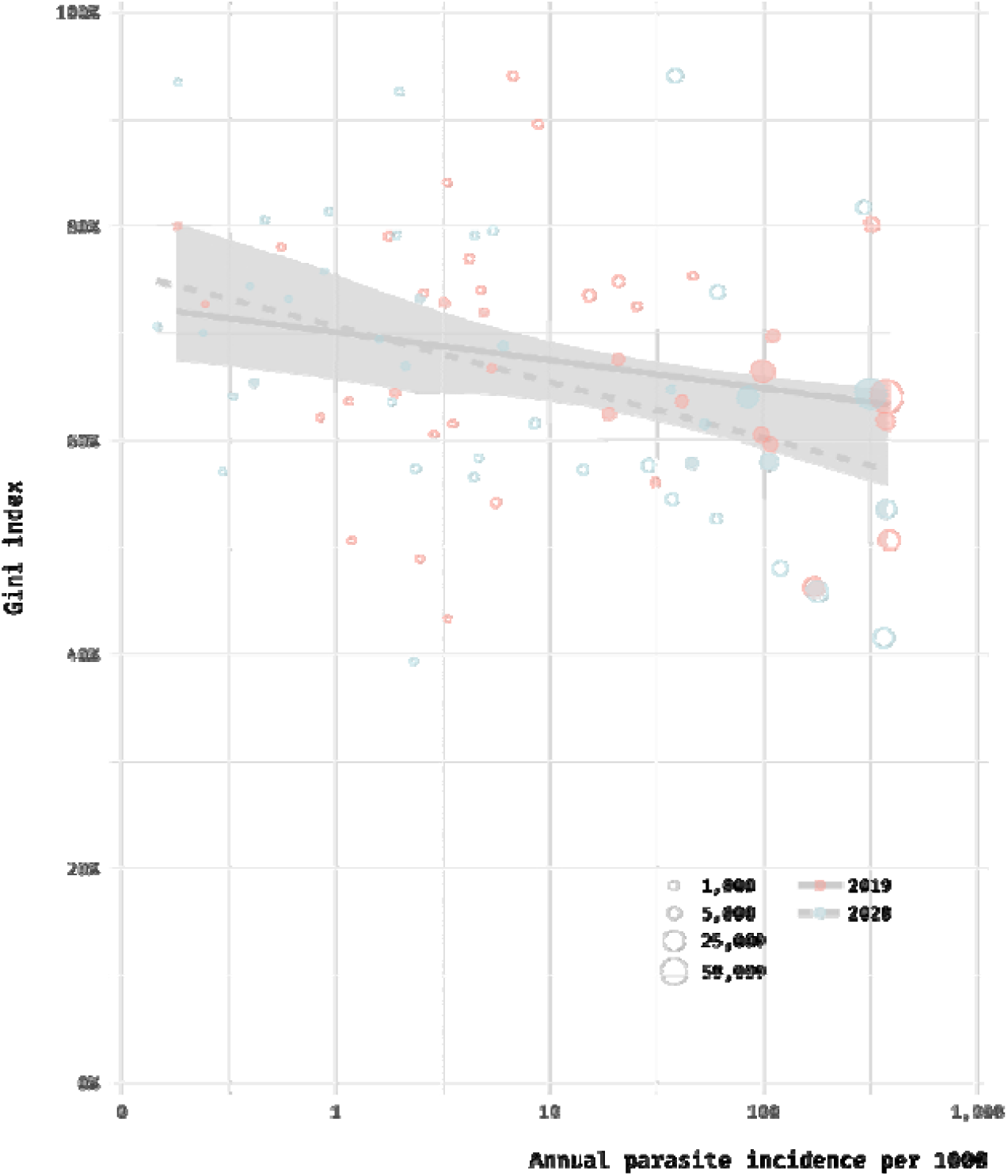
District-specific transmission intensity and its association with the Gini index. Straight lines were modelled by linear regression. Shaded grey areas around the lines represent the 95% confidence intervals. Each dot is proportional in size to the total number of cases. Horizontal axes are represented by a logarithmic scale.

## Discussion

### Key findings

Using large-scale, contemporary malaria surveillance data, this study aimed to quantify and characterise spatial heterogeneity in malaria transmission-intensity across Indonesian Papua. We observed that, between 2019 and 2020, the distribution of malaria across the region was highly heterogeneous. Our analysis indicates that this marked variation in transmission intensities was also linked to unique epidemiological characteristics of malaria in different areas.

We found heterogeneity of transmission ranging from high to extremely high (Gini index of [0.5, 1]). High Gini index values suggest the existence of a small number of spatial units or clusters (in our context, districts and health units) that experience a disproportionately greater burden of malaria than their counterparts. When we examined the data at the smaller spatial scale of health unit, we observed more pronounced heterogeneity, a common pattern with what had been reported from other countries.^16,20^ Not only within provinces, but also within districts, was there heterogeneity. We found noticeable variation in the within-district Gini index, suggesting there was clustering of the health facilities that accounted for a major proportion of cases despite containing a minority of the district’s population. Our findings emphasise the critical importance of not overlooking spatial heterogeneity at lower spatial resolution. Targeted interventions towards clustering cases at a lower level may improve efficiency and effectiveness because control efforts can be more focused. Risk factors in subpopulations of a particular cluster are likely to be more similar, allowing resource deployment with less operational complexity.

While conventional strategies for case reduction can be maintained or further scaled up in areas with a high malaria burden (such as in Keerom, where children were most clinically affected), more complex strategies to further shrink malaria clusters in low transmission settings that involve distinct transmission processes (such as in Jayawijaya, where transmission events peaked among older children and adults of working age) are equally essential for local elimination. Malaria elimination requires at least a three-year consecutive period of zero malaria incidence of indigenous origin, as well as fully functional surveillance and response system.^21^ Despite rational arguments that there is no one-size-fits-all path to elimination,^16^ malaria-eliminating countries have identified common features of regions on closely reaching elimination, including geospatial clustering and increased proportions of cases caused by *P. vivax*, as well as in males, imported, among migrants and marginalised populations.^22^ For instance, within the Brazilian Amazon Basin, high-transmission municipalities had cases predominantly among children and had similar proportions of men and women contracting malaria and while in low-transmission and near-elimination settings, the disease was more prevalent among men of working age.^16^

In our analysis, many districts with less intense transmission exhibited some of these features, where districts with lower API tended to have a higher proportion of cases attributable to *P. vivax* and occurring in adult men. Furthermore, lower transmission intensity was also associated with higher heterogeneity. Considering some similarities (e.g., malaria were broadly more prevalent among men than women) and differences (e.g., *P. vivax* had been long predominated in Brazil due to the more rapid decrease in *P. falciparum* cases under similarly high monthly incidence to Indonesian Papua, where *P. falciparum* still often slightly predominated or was equally prevalent to *P. vivax*), we observed comparable trends between the Amazon Basin and Indonesian Papua.^16^ That is, it suggests more generally that as a local area approaches local malaria elimination, we may expect more clustering and these clusters may demonstrate distinctive characteristics that are harder to control. For instance, *P. vivax* infection is known to be less susceptible to malaria control and elimination owing to, among other factors, its pre-onset gametocytaemia, more efficient transmission to vectors at lower parasitaemia, lower sensitivity of diagnostic tests for parasitaemia detection, dormant liver stages resulting in relapses and ongoing transmission, and poor adherence of patients to hypnozoiticidal treatment, which frequently involves a 14-day regimen associated with severe drug-induced haemolysis.^23^ These facts highlight the current challenges confronting locations with low transmission, which are mainly combating *P. vivax*, as well as the potential hurdle for locations in the Papua region, where both *P. falciparum* and *P. vivax* extensively coexist, as *P. vivax* may eventually be the ultimate nemesis before achieving elimination.^22^ A healthcare and surveillance system that is less sensitive to detecting and adapting to the region’s differentially changing malaria epidemiology could stall progress towards elimination.

According to mathematical models, the proportion of clinical malaria among under-5 children varies from over 60% to less than 20% in high- and low-transmission settings, respectively.^24^ Under-5 and younger children are also at considerable risk of morbidity and mortality^25^, and districts that continue to bear a disproportionate share of burden may prioritise reducing morbidity and mortality in this subpopulation. Meanwhile, districts with a case predilection towards male adults may focus on decreasing infection transmission.^22^ Differential age distributions between areas may also be indicative of infection transmission processes. Lana and colleagues suggest that because malaria affects all age and sex groups in high-transmission Brazilian municipalities, the infection distribution reflect the underlying demographic population.^16^ Transmission processes in these areas are typically peri-domestic in nature, i.e., occurring in or near human households.^16^ Areas in low intensity, on the other hand, may show a predisposition in men of working age^16^ that may be related to their occupation and behaviours outside the home.^22^

We observed that in many districts, a substantial proportion of malaria patients identified themselves as a farmer or a housewife, indicating transmission likely occurring in human settlements and agricultural fields. Bionomics of the primary vectors for malaria in Indonesian Papua, i.e., the *Anopheles punctulatus* complex, reflects the finding.^26^ These mosquitoes prefer open, sunlit spaces, either natural or artificial.^26^ This is in contrast to bionomics of the primary vectors in the adjacent Greater Mekong Subregion within Southeast Asia where most malarious regions are located in the forests (hence the term ‘forest malaria’) that involve more shaded spots and water collections for larval breeding.^27^ Greater understanding of this distinctive feature of vector bionomics may help in designing the most effective vector control in Indonesian Papua. Control measures that are effective in preventing peri-domestic transmission, such as insecticide-treated nets (emphasised heavily in the Papua region) but implemented in occupational and outdoor-related settings are unlikely to work. As in Brazil,^16^ human activities such as deforestation and migration likely played an important role in driving transmission heterogeneity and characteristics in Indonesian Papua. In 1980s, people transmigration from more populated islands of Java, Lombok, and Bali was initiated to newly deforested locations across the endemic Papua region.^28^ This in part led to the abundance of breeding sites for the *An. punctulatus* complex and mixing of nonimmune and malaria-carrying individuals.^28^

Recently, a phase-2b R21/MM malaria vaccine randomised trial conducted in Burkina Faso showed that the vaccine was more than 75% effective at reducing the risk of clinical falciparum malaria in children aged 5–17 months.^29^ While this is encouraging, particularly for populations where *P. falciparum* malaria still predominates in young children (e.g., districts with the highest intensity such as Keerom and Sarmi), the relevance of vaccines designed to mitigate early childhood mortality in African settings may be less impactful in settings such as Indonesian Papua. Vaccines that increase the pool of asymptomatic carriers may also not be suitable for low- and near-elimination settings where this pool is likely the main problem.

Over the last decade, the cumulative incidence of malaria in the Papua region had fallen dramatically, owing, in part, to increased funding and capacity building for malaria control involving improved vector control, case detection, antimalarial therapy, and strengthened health systems, as well as socio-economic development.^7^ These factors may have acted differentially across the region. In particular, endemicity in West Papua province, where a monthly incidence risk of fewer than two cases per 1,000 population, had been sustained since late 2015. Papua province, on the other hand, had since experienced a rising trajectory. Districts with the highest API (e.g., of [100, 1000) cases per 1000 population) remained restricted to Papua province, while districts with a lower baseline API in 2019 tended to achieve a greater relative reduction in API in the following year, further amplifying transmission heterogeneity.

### Strengths and limitations

The strengths of this study include its extensive use of large-scale and contemporary data derived from the routinely collected malaria surveillance system in Indonesia. These data, which cover over 446,000 malaria patients in the past two years alone, incorporate cases identified through a variety of methods, including passive case detection, house visits, active mass blood surveys, and prenatal screening. This breadth and depth of data may enable us to improve estimation accuracy and precision, either overall or within subpopulations of interest. Second, we quantified spatial heterogeneity using the Gini index. This index has been shown to be a sensitive and intuitive measure to capture the heterogeneity of transmission intensity over space and time.^15^ When compared to other potential measures of heterogeneity, such as the coefficient of variation, which can exceed 100% while being highly correlated with the Gini index, the Gini index, bounded between [0, 100]%, can facilitate interpretation and comparability across studies.^15^ Along with visualising different levels of malaria intensity using geospatial maps, summarising heterogeneity into a single metric using the Gini index may be useful and complementary for tracking progress towards elimination.^15^

Several aspects may limit our study. At the moment, we can only estimate the Gini index in 2019 and 2020, obscuring the trend of heterogeneity over time. In Brazil, heterogeneity increased in a roughly linear manner over time as malaria incidence declined, irrespective of administrative levels considered.^16^ However, because the Sismal database is continually updated, our study can serve as a starting point for research on temporal dynamics of spatial heterogeneity. Furthermore, using districts as observation units (instead of years), we did find an approximately linear, inverse relationship between transmission intensity and spatial heterogeneity. We still lacked information on several important characteristics of notified malaria cases, such as infection sources (indigenous or imported) and recurrence status (reinfection, relapse, or recrudescence). In the current version of the database, several of these features either had a high proportion of missingness or were recorded in an uninformative way. Given that the database is mostly (90% or more) composed of routinely collected data from healthcare services, it is reasonable to expect that the improvements in collecting information from notified cases in the clinics would benefit the surveillance system and research purposes.^30^

### Implications

Malaria remains a serious problem that impacts people of all ages and various occupations in Indonesian Papua because transmission occurs where they live, work, and go to school. That transmission is highly heterogeneous and unique in its epidemiological features. This emphasises that a one-size-fits-all approach to malaria control and elimination is unlikely to be the most cost-effective strategy, and that additional attention to spatially explicit interventions may be required. Distinct epidemiological characteristics also imply that more tailored resource priorities may be needed according to each area’s transmission level. By leveraging routinely collected data across the region and the Gini index to measure heterogeneity, stakeholders now have a method for identifying and characterising malaria transmission hotspots on a regular basis, allowing interventions to be targeted to those most vulnerable. Locally enhanced epidemiologic investigations and surveillance can be extremely invaluable, for instance, by increasing the capacity of malaria village workers. Lastly, we noted several opportunities to further improve the data quality of the malaria surveillance system for research purposes.

## Data Availability

The datasets supporting the conclusions of this study can be accessed at http://www.sismal.malaria.id/ (available upon request to the Indonesian Sub-Directorate for Malaria Control), https://www.bps.go.id/ (publicly available), and http://bppsdmk.kemkes.go.id/web/ (publicly available) for the individual-level malaria surveillance data, the projected population sizes, and the numbers of health units in the Papua region, respectively.

## Contributors

IF and IRFE conceived the study. GA, SBF, ES, BGM, and BW were responsible for data collection and provision. IF, BAD, KDL, and IRFE curated and verified the data. IF was responsible for formal analysis, visualisation, and writing the initial draft with input from BAD, HS, and IRFE. IM, JM, JKB, and IRFE were responsible for project oversight. All authors had access to the data, critically reviewed the study for intellectual content, and had full responsibility for the decision to submit the final manuscript.

## Data sharing

The datasets supporting the conclusions of this study can be accessed at http://www.sismal.malaria.id/ (available upon request to the Indonesian Sub-Directorate for Malaria Control), https://www.bps.go.id/ (publicly available), and http://bppsdmk.kemkes.go.id/web/ (publicly available) for the individual-level malaria surveillance data, the projected population sizes, and the numbers of health units in the Papua region, respectively. Reproducible code for data analysis and visualisation is available at https://github.com/ihsanfadil/spark-hetero.

## Acknowledgments

This study was funded by the Centre for Health Security at the Australian Department of Foreign Affairs and Trade through the Strengthening Preparedness in the Asia-Pacific Region through Knowledge (SPARK) project. We are grateful to all the healthcare staff who contributed to the establishment and maintenance of the malaria surveillance system, particularly the provincial and district health offices in Papua and West Papua, as well as the Sub-Directorate for Malaria Control, for making the data accessible. We thank Dr Narimane Nekkab who helped clarify the mathematical formula of the Gini index and its R-code implementation.

