## Supplementary Material for "Quantifying spatial heterogeneity of malaria in the endemic Papua region of Indonesia: analysis of epidemiological surveillance data"

**Figure S1. Spatial heterogeneity of transmission intensity among the districts in 2019 and 2020**. Each data point represents the district-specific annual parasite incidence. Horizontal axis is shown on the logarithmic scale.

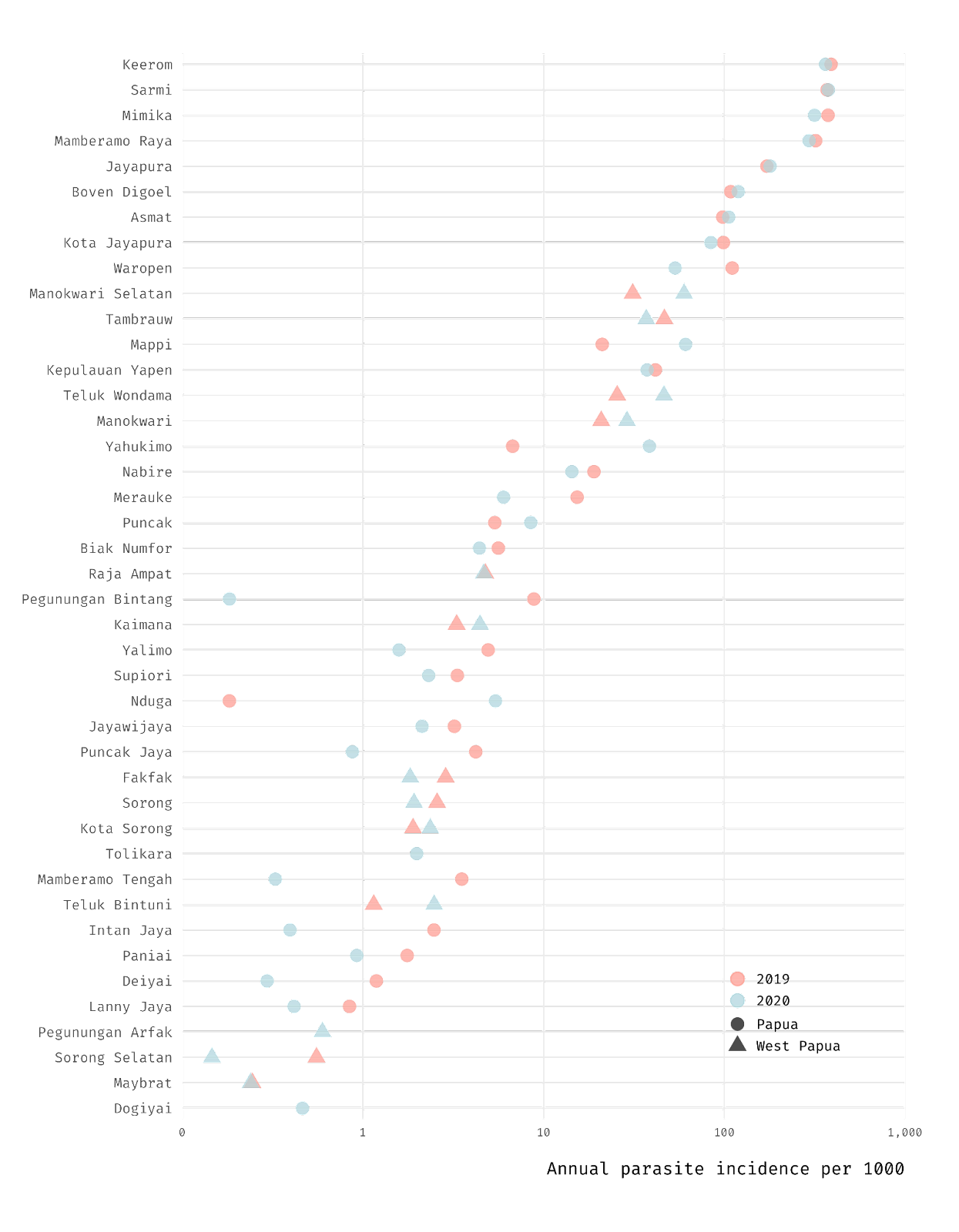

**Figure S2. Distributions of malaria cases by sex and parasite species in Keerom, Nabire, and Jayawijaya**. Vertical axis denotes the monthly incidence risk of malaria per 1,000 population in 2020, stratified by sex and parasite species. Each plot represents a district with a relatively high transmission intensity (Keerom), a moderate transmission intensity (Nabire), or a low transmission intensity (Jayawijaya).

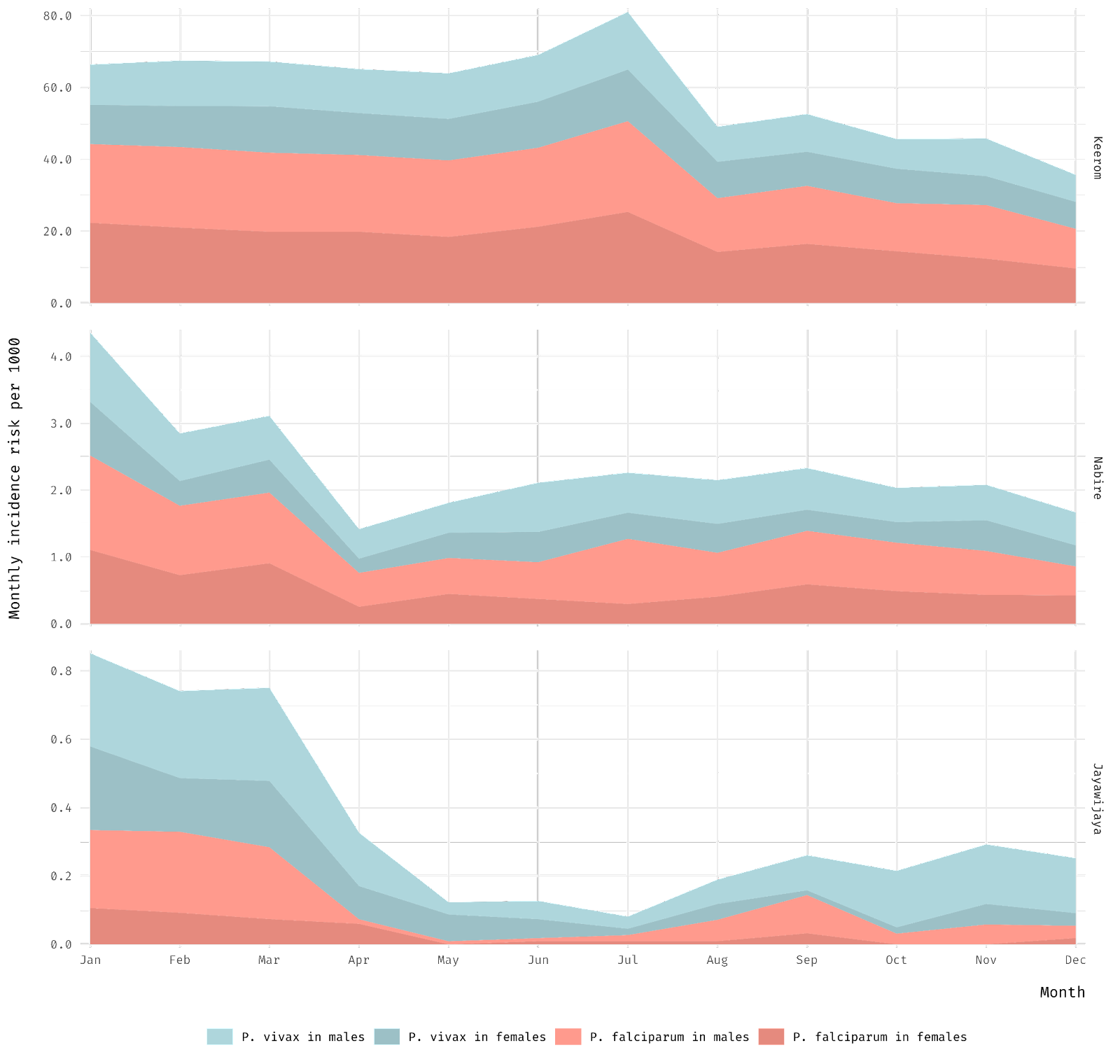

**Figure S3. Relative proportions of clinical severity of malaria cases in different health-unit types, by year and province**. In general, health centres and clinics provide primary care, whereas hospitals admit referrals from primary care, as well as more severe and/or complicated cases requiring specialist physicians. NA not available.

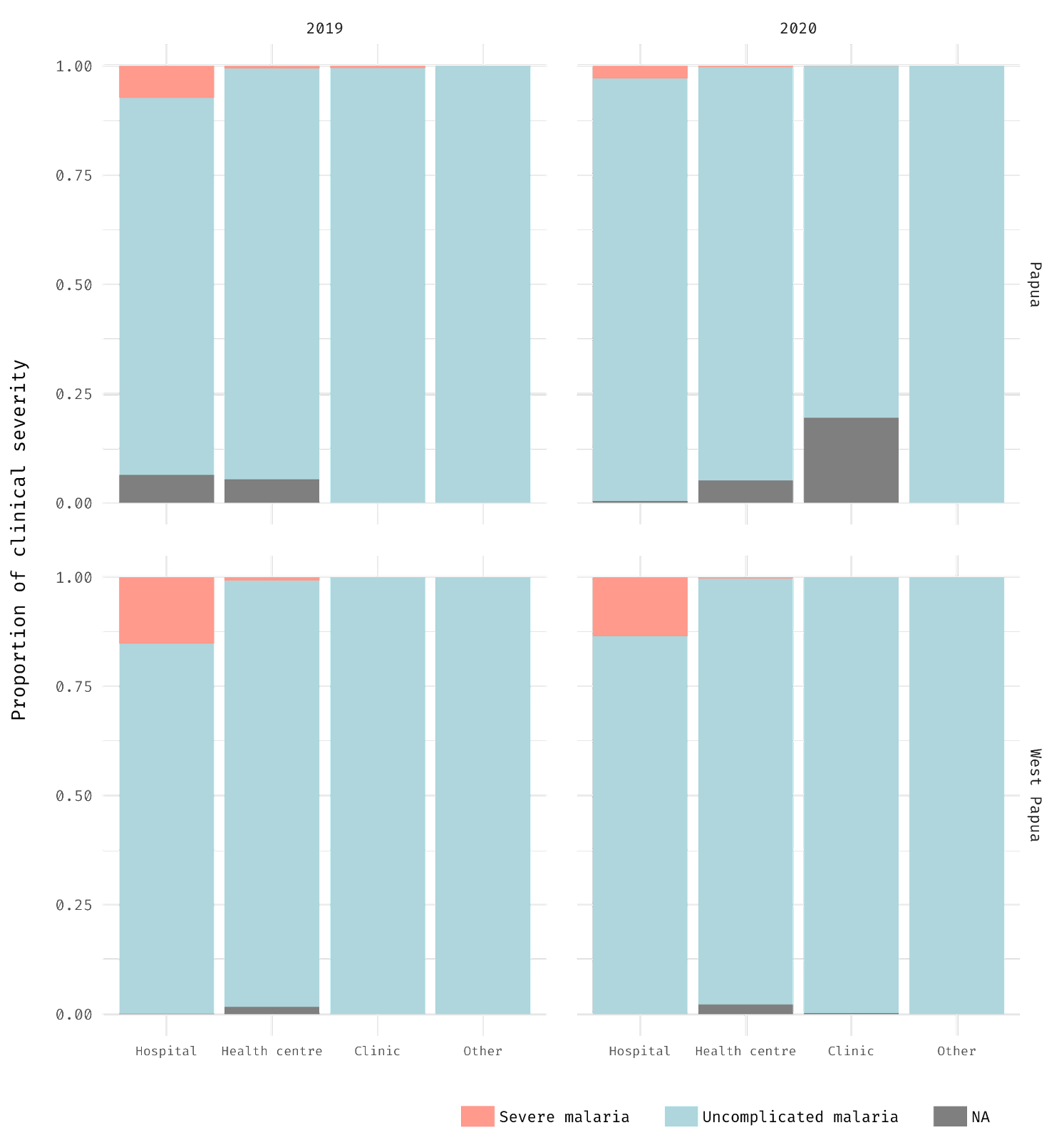

**Figure S4. District-specific proportions of health unit types that detected malaria cases, by year and province**. (A) Papua. (B) West Papua.

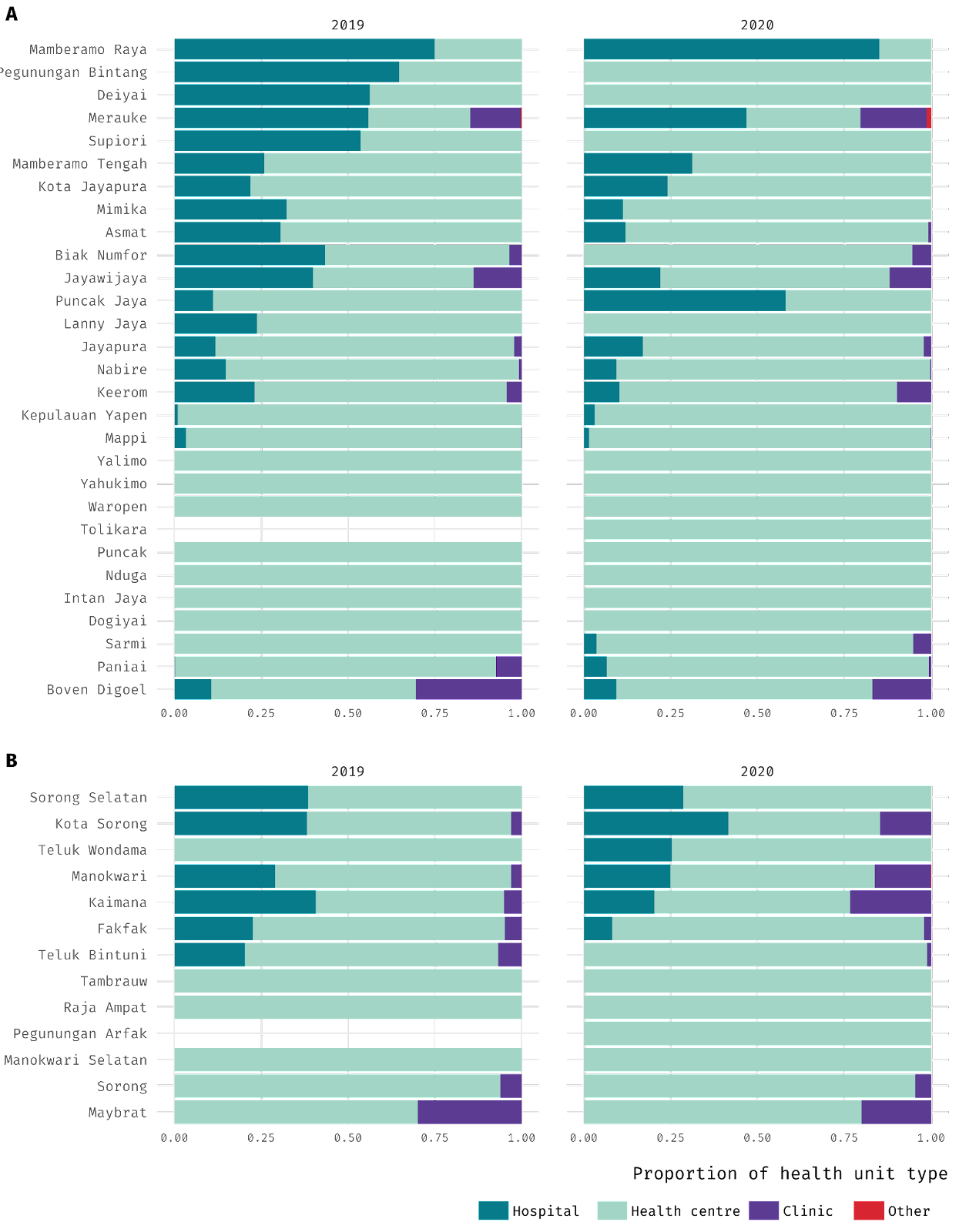

**Figure S5. Full bootstrap distributions of the Gini index estimates evaluated at the district level.** 2·5th and 97·5th percentile values represent the lower and upper limits of the confidence interval. A Gini index of zero and one are equivalent to a value of 0% and 100%, respectively.

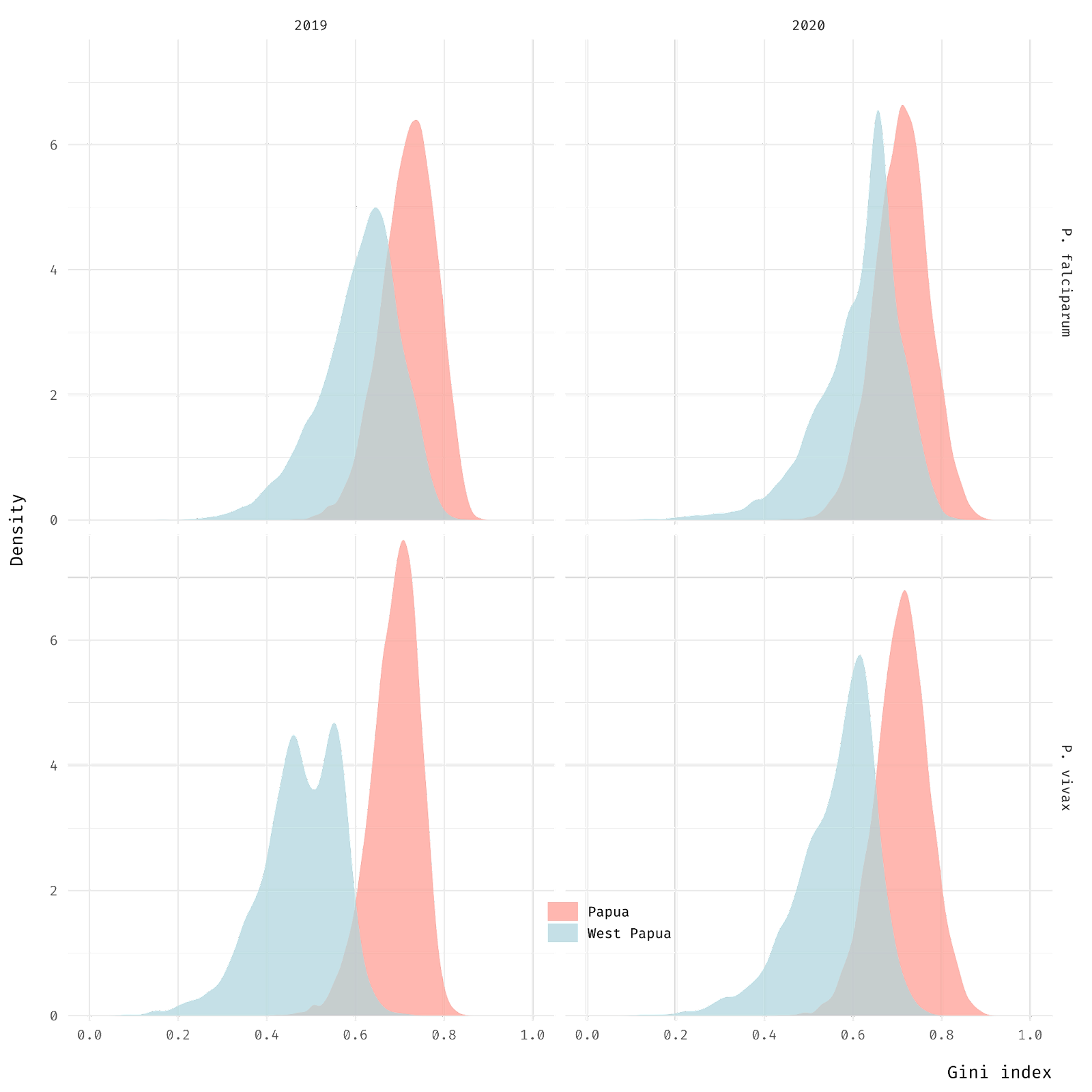

**Figure S6. Full bootstrap distributions of the Gini index estimates evaluated at the health unit level**. 2·5th and 97·5th percentile values represent the lower and upper limits of the confidence interval. A Gini index of zero and one are equivalent to a value of 0% and 100%, respectively.

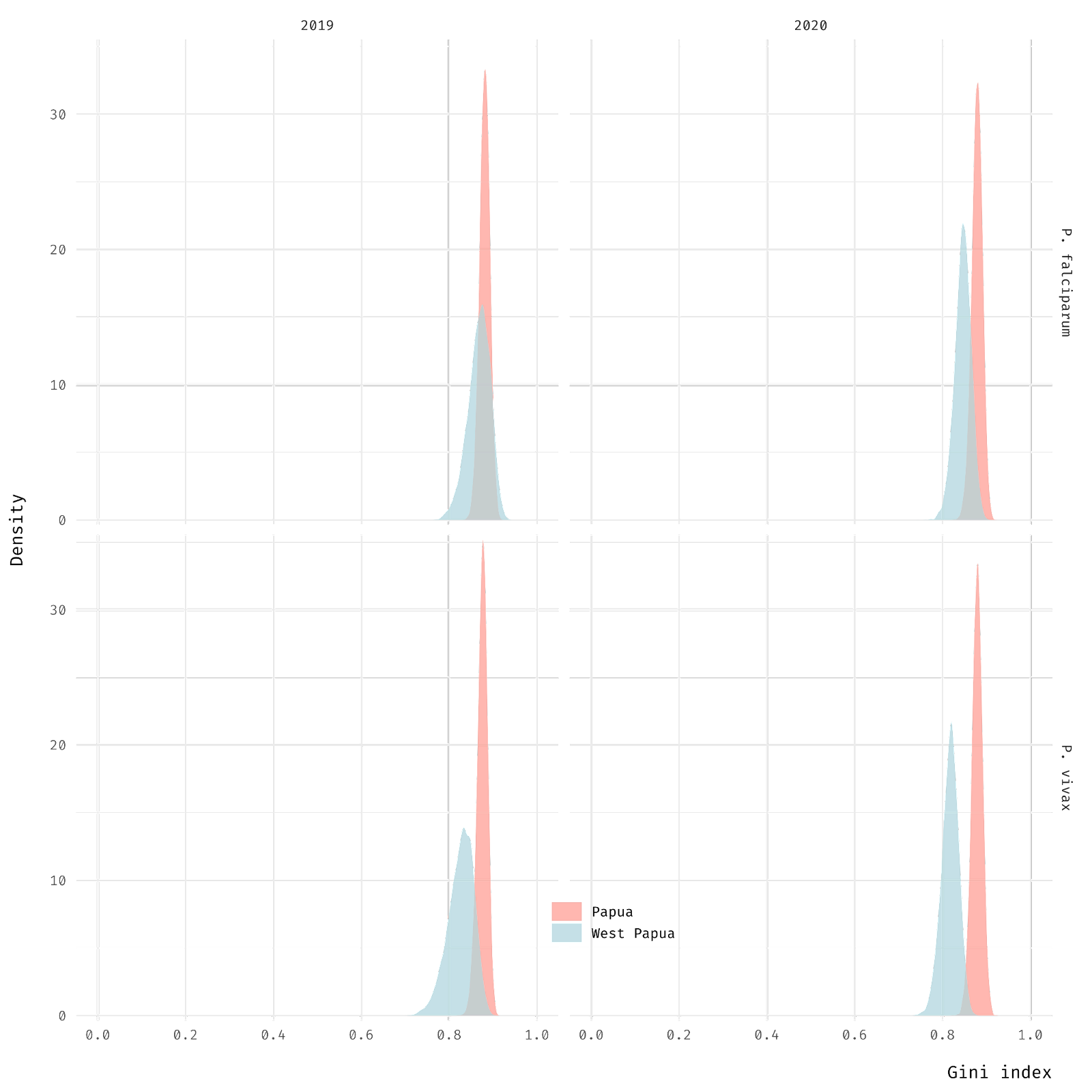
